# Association Between Childhood Autism and Speech Development Difficulties: A Retrospective Cross-Sectional Study From Tajikistan

**DOI:** 10.64898/2026.02.01.26345313

**Authors:** Mirsafo Mirsharofov

## Abstract

**Background:** Autism spectrum disorder (ASD) is frequently associated with speech and language difficulties, yet empirical data from Central Asian countries remain scarce. This study examined the association between a diagnosis of childhood autism (ICD-10: F84.0) and the presence of speech development difficulties in a clinical sample from Tajikistan

**Method:** A retrospective cross-sectional study was conducted using clinical records of 85 patients (36 with F84.0; 49 with other psychiatric diagnoses) at the Insight Mental Health Center in Dushanbe, Tajikistan (December 2025–January 2026). Speech difficulties were identified through systematic review of clinical notes. Between-group comparisons were performed using Pearson’s χ^2^ test, odds ratios (OR), relative risk (RR), and effect size measures (φ coefficient, Cohen’s h).

**Results:** Speech difficulties were present in 72.2% of the autism group versus 36.7% of the comparison group. The association was statistically significant (χ^2^ = 10.47, p <.01). Children with autism had substantially higher odds of speech difficulties (OR = 4.48, 95% CI [1.76, 11.38]), with a large effect size (Cohen’s h = 0.73).

**Conclusions:** Autism diagnosis was significantly associated with elevated rates of speech difficulties in this Tajik clinical sample.

**Practical implications:** These findings support the systematic inclusion of speech-language assessment and intervention within autism care protocols, particularly in Central Asian healthcare settings where such integration remains limited.

**Highlights:**

- Speech difficulties were identified in 72.2% of children with autism (F84.0) in a Tajik clinical sample.
- Children with autism were 4.5 times more likely to present with speech difficulties than those with other diagnoses (OR = 4.48, 95% CI [1.76, 11.38]).
- The most prevalent speech pattern was complete absence of expressive speech (nonverbal presentation).
- Findings support the integration of speech-language assessment into standard autism care protocols in Central Asia.
- This is one of the first empirical reports on autism and speech profiles from Tajikistan.

## 1. Introduction

Autism spectrum disorder (ASD) is a neurodevelopmental condition characterized by persistent differences in social communication and social interaction, along with restricted, repetitive patterns of behavior, interests, or activities (American Psychiatric Association [APA], 2013). Global prevalence estimates indicate that approximately 1 in 100 children are autistic (World Health Organization [WHO], 2023), though reported rates vary considerably by region and methodology (Zeidan et al., 2022).

Speech and language difficulties occupy a central position in the clinical profile of autism. International research has consistently demonstrated that between 25% and 50% of autistic children remain minimally verbal or nonspeaking by school age (Tager-Flusberg & Kasari, 2013). The spectrum of speech-related challenges in autism is broad, encompassing the complete absence of expressive speech, echolalia, stereotyped utterances, pragmatic language differences, and regression of previously acquired language skills (Boucher, 2012; Kjelgaard & Tager-Flusberg, 2001). Longitudinal evidence suggests that language trajectories in autism are highly heterogeneous, with some children showing significant gains through childhood and adolescence and others remaining nonspeaking (Pickles et al., 2014).

Despite the well-established link between autism and language difficulties in Western research contexts, empirical data from Central Asian countries remain remarkably scarce. A recent systematic review of global autism research output found that the vast majority of published studies originate from the United States, the United Kingdom, Australia, and Canada, with low- and middle-income countries, particularly those in Central Asia, being severely underrepresented (Hahler & Elsabbagh, 2015; Memisevic et al., 2023). This geographic imbalance is problematic because the clinical presentation, identification, and management of autism may be shaped by cultural, linguistic, and healthcare system factors that differ substantially across regions (Durkin et al., 2015; Elsabbagh et al., 2012).

In Tajikistan, a lower-middle-income country in Central Asia with a population of approximately 10 million, the infrastructure for diagnosing and supporting autistic individuals remains limited. There is a scarcity of specialized professionals, standardized diagnostic tools have not been formally validated in the Tajik or Russian languages as used locally, and systematic epidemiological data on autism prevalence are unavailable. Under these circumstances, understanding the clinical characteristics of autistic children who present to specialized centers is an important first step in developing evidence-based service provision.

The Insight Mental Health Center in Dushanbe is one of the few facilities in Tajikistan providing specialized assessment and intervention for children with neurodevelopmental conditions, including autism. Preliminary analysis of the center’s clinical database, facilitated by artificial intelligence (AI)-assisted data mining, revealed that speech-related symptoms were among the most frequently documented clinical features, appearing in records associated with 62 patients across 128 clinical entries. This observation motivated a formal examination of the association between autism diagnosis and speech difficulties.

### 1.1. Aims and Hypotheses

The primary aim of this study was to examine the association between a diagnosis of childhood autism (ICD-10: F84.0; WHO, 2019) and the presence of speech development difficulties in a clinical sample from Tajikistan. We hypothesized that patients with a diagnosis of F84.0 would demonstrate a significantly higher prevalence of speech difficulties compared with patients with other psychiatric diagnoses. A secondary aim was to describe the types of speech difficulties observed in the autism group. This study contributes to the growing body of literature on autism in underrepresented global contexts and has direct practical implications for service development in Central Asian healthcare settings.

## 2. Method

### 2.1. Study Design and Setting

This was a retrospective cross-sectional study conducted at the Insight Mental Health Center, a specialized outpatient facility in Dushanbe, Tajikistan. The center provides psychiatric assessment, diagnosis, and intervention services for children and adults with a range of mental health conditions. Ethical approval for this retrospective analysis of anonymized clinical data was obtained from the institutional review board of the Insight Mental Health Center. The study was conducted in accordance with the ethical principles of the Declaration of Helsinki.

### 2.2. Participants

Clinical records of 85 unique patients who attended the Insight Mental Health Center between December 2025 and January 2026 were included. Inclusion criteria were: (a) an established psychiatric diagnosis according to ICD-10, and (b) availability of clinical documentation including presenting complaints and clinical observations. The sole exclusion criterion was the absence of clinical data in the electronic record.

Patients were allocated to two groups. The autism group (n = 36) comprised patients with a primary or secondary diagnosis of childhood autism (F84.0). The comparison group (n = 49) included patients with other psychiatric diagnoses, including hyperkinetic disorder (F90.0), mild intellectual disability (F71), specific developmental disorders of speech and language (F80.x), depressive episodes (F32.x), anxiety disorders (F41.x), obsessive-compulsive disorder (F42), and schizophrenia (F20.x), among others.

### 2.3. Measures

#### 2.3.1. Autism Diagnosis

Diagnoses of childhood autism (F84.0) were established by qualified psychiatrists following clinical assessment in accordance with ICD-10 diagnostic criteria (WHO, 2019). While structured diagnostic instruments such as the Autism Diagnostic Observation Schedule (ADOS-2; Lord et al., 2012) and the Autism Diagnostic Interview-Revised (ADI-R; Rutter et al., 2003) are not yet routinely available in Tajikistan, clinical diagnoses were made by experienced clinicians using comprehensive developmental history, behavioral observation, and collateral information from caregivers.

#### 2.3.2. Speech Difficulties

The presence of speech difficulties was determined through a systematic review of clinical records, including documented presenting complaints, objective clinical observations, and notes from therapeutic sessions. A patient was classified as having speech difficulties if any of the following features were documented: complete absence of expressive speech (nonspeaking presentation); minimal speech (fewer than 10 words); echolalia (immediate or delayed); presence of vocalizations only without formed words; disordered or incomplete speech with impaired intelligibility; articulation difficulties (dyslalia); or regression of previously acquired speech skills. This classification was performed by the first author, a qualified psychiatrist with expertise in child mental health and autism.

### 2.4. Statistical Analysis

Between-group differences in the frequency of speech difficulties were analyzed using Pearson’s chi-square (χ^2^) test with Yates’ correction. The strength of association was quantified using the phi (φ) coefficient, odds ratio (OR) with 95% confidence interval (Woolf method), and relative risk (RR). Effect size was additionally evaluated using Cohen’s h. The threshold for statistical significance was set at p <.05. All analyses were performed using a purpose-built analytical system integrated into the center’s clinical database.

## 3. Results

### 3.1. Sample Characteristics

The total sample comprised 85 patients. The autism group (n = 36) consisted of 27 males (75.0%) and 9 females (25.0%), yielding a male-to-female ratio of 3.0:1. The mean age in the autism group was 5.8 years (range: 2–12 years; median: 6 years); all participants in this group were children. The comparison group (n = 49) was more heterogeneous, comprising 20 children and 29 adults with a range of psychiatric diagnoses.

### 3.2. Prevalence of Speech Difficulties

Speech difficulties were documented for 44 of 85 patients in the total sample (51.8%).

Within the autism group, speech difficulties were present in 26 of 36 patients (72.2%), compared with 18 of 49 patients (36.7%) in the comparison group. The between-group difference was 35.5 percentage points. Table 1 presents the contingency table for the association between autism diagnosis and speech difficulties.

**Table 1.** Contingency Table: Autism Diagnosis (F84.0) and Speech Difficulties.

| Group | Speech (+) | Speech (–) | Total | % Speech (+) |
| --- | --- | --- | --- | --- |
| F84.0 (+) | 26 | 10 | 36 | 72.2 |
| F84.0 (–) | 18 | 31 | 49 | 36.7 |
| **Total** | **44** | **41** | **85** | **51.8** |

### 3.3. Statistical Analysis

The Pearson χ^2^ test revealed a statistically significant association between autism diagnosis and speech difficulties (χ^2^ = 10.47, df = 1, p <.01). This result was confirmed with Yates’ correction (χ^2^ = 9.09, p <.01). Table 2 summarizes the full results of the statistical analysis.

**Table 2.** Results of Statistical Analysis.

| Statistic | Value | Interpretation |
| --- | --- | --- |
| Pearson $\chi^2$ | 10.47 | $p < .01$ |
| $\chi^2$ (Yates' correction) | 9.09 | $p < .01$ |
| Phi ($\phi$) coefficient | 0.351 | Medium effect |
| Odds ratio (OR) | 4.48 | Strong association |
| 95% CI for OR | [1.76, 11.38] | Significant |
| Relative risk (RR) | 1.97 | Nearly 2-fold increase |
| Cohen's $h$ | 0.73 | Large effect |

The odds ratio of 4.48 indicates that patients with childhood autism had approximately 4.5 times the odds of presenting with speech difficulties compared with patients carrying other diagnoses. The lower bound of the 95% confidence interval (1.76) exceeds 1.0, confirming the statistical significance of this association. The relative risk of 1.97 indicates that the probability of speech difficulties was nearly twice as high in the autism group. The phi coefficient of 0.351 indicates a medium-strength association, while Cohen’s h of 0.73 represents a large effect size, underscoring the clinical significance of the finding.

### 3.4. Types of Speech Difficulties in the Autism Group

Among the 26 patients in the autism group who presented with speech difficulties, the following patterns were observed: complete absence of expressive speech (nonspeaking presentation) was the most prevalent pattern; minimal speech limited to 1–6 words; echolalia (immediate and delayed); vocalizations without formed words; disordered and incomplete speech with reduced intelligibility; and regression of previously acquired speech skills. Due to the retrospective nature of the data and the absence of standardized speech-language assessments, precise frequencies for each category could not be reliably quantified.

## 4. Discussion

This study examined the association between a diagnosis of childhood autism (F84.0) and speech development difficulties in a clinical sample from Tajikistan. The results confirmed our hypothesis: children with autism were significantly more likely to present with speech difficulties than patients with other psychiatric diagnoses. The prevalence of speech difficulties in the autism group (72.2%) and the large effect size (Cohen’s h = 0.73) underscore the clinical importance of this association.

### 4.1. Comparison With International Literature

The observed rate of speech difficulties among autistic children (72.2%) is broadly consistent with, though somewhat higher than, estimates from international studies. Tager-Flusberg and Kasari (2013) reported that 25–50% of autistic children remain minimally verbal by school age, while Rose et al. (2016) found that approximately 30–40% of children in community-based early intervention programs were minimally verbal. The somewhat higher rate in the present study may reflect several factors. First, children presenting to a specialized mental health center may have more pronounced clinical features than those identified in community-based samples. Second, the inclusion of a broad range of speech-related challenges—from minimal speech to echolalia and articulation difficulties—likely captured a wider spectrum of difficulties than studies focused exclusively on minimally verbal or nonspeaking children. Third, the absence of accessible early intervention services in Tajikistan may result in children presenting at later ages with more entrenched speech difficulties, as suggested by the mean age at presentation of 5.8 years in this sample.

The male-to-female ratio of 3.0:1 in the autism group aligns well with the meta-analytic estimate of approximately 3–4:1 reported by Loomes et al. (2017). The mean age of presentation (5.8 years), however, is notably later than recommended by international guidelines, which advocate for autism screening at 18 and 24 months (Hyman et al., 2020). This late identification pattern is consistent with findings from other low- and middle-income countries (Durkin et al., 2015) and highlights the need for improved early detection systems in Tajikistan.

### 4.2. Practical Implications

The findings have several practical implications for clinical practice and service development in Tajikistan and potentially other Central Asian countries. First, the high prevalence of speech difficulties among autistic children supports the systematic inclusion of speech-language assessment as part of the standard evaluation protocol when autism is diagnosed or suspected. At present, access to speech-language pathology services in Tajikistan is limited, and formal integration of these services into autism care pathways has not been established.

Second, the predominance of nonspeaking presentations among autistic children underscores the need for training in augmentative and alternative communication (AAC) approaches, such as the Picture Exchange Communication System (PECS; Bondy & Frost, 1994) or speech-generating devices. These approaches have demonstrated efficacy in supporting communication development in autistic children who are nonspeaking or minimally verbal (Ganz et al., 2012) and could be adapted for use in Tajik clinical settings.

Third, the late age at presentation highlights the urgent need for community-based early screening programs. The Modified Checklist for Autism in Toddlers, Revised with Follow-Up (M-CHAT-R/F; Robins et al., 2014) has been successfully implemented in diverse cultural contexts and could be considered for adaptation and validation in Tajikistan. Early identification would enable earlier access to speech-language intervention, which is associated with better long-term communication outcomes (Rogers et al., 2012).

### 4.3. Limitations

Several limitations should be considered when interpreting these findings. First, the retrospective design precludes causal inference; the observed association does not establish that autism causes speech difficulties, as both may share underlying neurodevelopmental origins.

Second, the sample was drawn from a single clinical center, which limits generalizability to the broader population of autistic individuals in Tajikistan. Third, the comparison group was heterogeneous, including both children and adults with a range of diagnoses, which may have influenced the between-group comparison. A more tightly matched comparison group of children with non-autism neurodevelopmental conditions would strengthen future analyses.

Fourth, the identification of speech difficulties relied on review of clinical records rather than standardized speech-language assessments. While systematic criteria were applied, the absence of validated instruments such as the Clinical Evaluation of Language Fundamentals (CELF-5; Wiig et al., 2013) or the Preschool Language Scales (PLS-5; Zimmerman et al., 2011) introduces potential measurement variability. Fifth, the relatively small sample size (N = 85) limits statistical power and the precision of effect estimates, as reflected in the wide confidence interval for the odds ratio. Finally, autism diagnoses were based on clinical judgment rather than structured diagnostic instruments (e.g., ADOS-2, ADI-R), which is a common limitation in resource-constrained settings but may affect diagnostic specificity.

### 4.4. Future Directions

Future research should address these limitations through prospective, multi-center studies using standardized diagnostic and speech-language assessment tools. Of particular value would be studies examining predictors of speech development in autistic children in Central Asia, including the impact of intervention type, intensity, and age at initiation. Comparative effectiveness research on communication interventions (e.g., ABA-based approaches, PECS, TEACCH) adapted for Central Asian contexts would directly inform clinical practice.

Additionally, epidemiological studies are needed to establish the prevalence and characteristics of autism in Tajikistan, which would support the development of national screening and service delivery strategies.

## 5. Conclusion

This study provides one of the first empirical examinations of the association between autism and speech difficulties in a clinical sample from Tajikistan. Children diagnosed with childhood autism (F84.0) were significantly more likely to present with speech difficulties than patients with other psychiatric diagnoses, with a large effect size supporting the clinical relevance of this finding. The results highlight the critical importance of integrating speech-language assessment and intervention into standard autism care protocols in Tajikistan and support the development of early screening and intervention programs adapted to the Central Asian context. By contributing data from an underrepresented region, this study responds to calls for more globally diverse autism research and underscores the universal relevance of addressing communication needs in autistic individuals.

## Declarations

### Ethical Approval

Ethical approval for this retrospective analysis of anonymized clinical data was obtained from the institutional review board of the Insight Mental Health Center, Dushanbe, Tajikistan. The study was conducted in accordance with the Declaration of Helsinki.

### CRediT Authorship Contribution Statement

Mirsafo Bakaev: Conceptualization, Methodology, Data curation, Formal analysis, Investigation, Writing – original draft, Writing – review & editing.

### Data Availability

Data are available from the corresponding author upon reasonable request, subject to ethical and privacy considerations.

